# Propofol for treatment resistant depression: A randomized controlled trial

**DOI:** 10.1101/2023.09.12.23294678

**Authors:** Scott C. Tadler, Keith G. Jones, Carter Lybbert, Jason C. Huang, Rana Jawish, Daniela Solzbacher, E. Jeremy Kendrick, Matthew D. Pierson, Kamile Weischedel, Noreen Rana, Rebecca Jacobs, Lily C. Vonesh, Daniel A. Feldman, Claire Larson, Nathan Hoffman, Jacob E. Jessop, Adam L. Larson, Norman E. Taylor, David H. Odell, Kai Kuck, Brian J. Mickey

## Abstract

**Background:** Anesthetic agents including ketamine and nitrous oxide have shown antidepressant properties when appropriately dosed. Our recent open-label trial of propofol, an intravenous anesthetic known to elicit transient positive mood effects, suggested that it may also produce robust and durable antidepressant effects when administered at a high dose that elicits an electroencephalographic (EEG) burst-suppression state. Here we report findings from a randomized controlled trial (NCT03684447) that compared two doses of propofol. We hypothesized greater improvement with a high dose that evoked burst suppression versus a low dose that did not.

**Methods:** Participants with moderate-to-severe, treatment-resistant depression were randomized to a series of 6 treatments at low versus high dose (n=12 per group). Propofol infusions were guided by real-time processed frontal EEG to achieve predetermined pharmacodynamic criteria. The primary and secondary depression outcome measures were the 24-item Hamilton Depression Rating Scale (HDRS-24) and the Patient Health Questionnaire (PHQ-9), respectively. Secondary scales measured suicidal ideation, anxiety, functional impairment, and quality of life.

**Results:** Treatments were well tolerated and blinding procedures were effective. The mean [95%-CI] change in HDRS-24 score was −5.3 [−10.3, −0.2] for the low-dose group and −9.3 [−12.9, −5.6] for the high-dose group (17% versus 33% reduction). The between-group effect size (standardized mean difference) was −0.56 [−1.39, 0.28]. The group difference was not statistically significant (p=0.24, linear model). The mean change in PHQ-9 score was −2.0 [−3.9, −0.1] for the low dose and −4.8 [−7.7, −2.0] for the high dose. The between-group effect size was −0.73 [−1.59, 0.14] (p=0.09). Secondary outcomes favored the high dose (effect sizes magnitudes 0.1 - 0.9) but did not generally reach statistical significance (p>0.05).

**Conclusions:** The medium-sized effects observed between doses in this small, controlled, clinical trial suggest that propofol may have dose-dependent antidepressant effects. The findings also provide guidance for subsequent trials. A larger sample size and additional treatments in series are likely to enhance the ability to detect dose-dependent effects. Future work is warranted to investigate potential antidepressant mechanisms and dose optimization.

## Introduction

For over half a century, the effectiveness of psychopharmacological treatments remained essentially unchanged. No comparably efficacious medical alternatives to electroconvulsive therapy (ECT) existed. Despite its safety and efficacy, ECT has been relegated to use with the sickest, most treatment-resistant patients because of ECT-related side effects and public skepticism. More recently, however, newer and rapidly effective antidepressant and antisuicidal therapies have started to reshape the landscape. Most well-known is ketamine. Others showing promise and in various stages of investigation include nitrous oxide, isoflurane, propofol, and psychedelic/empathogenic compounds (e.g., psilocybin). Here we describe the first controlled trial using propofol anesthesia to treat depression.

Evidence has accumulated suggesting that some anesthetics and related psychoactive agents have antidepressant properties that long outlast their pharmacokinetic effects.^1,2^ Ketamine and nitrous oxide, which block the N-methyl-D-aspartate (NMDA) glutamate receptor, have antidepressant effects when administered at sub-anesthetic doses ^3–6^. The gas anesthetic isoflurane has shown antidepressant properties at high doses that produce EEG burst suppression ^7–10^, which consists of repeated bursts of activity separated by quiescent periods. ^11–15^. Isoflurane acts mainly as a positive allosteric modulator of the type-A γ-aminobutyric acid (GABA-A) receptor, but the NMDA receptor has also been implicated ^16,17^. Another GABA-A positive allosteric modulator, allopregnanolone (brexanolone), has antidepressant activity at sub-anesthetic doses and was recently approved for clinical use ^18^.

Propofol is an intravenous anesthetic that has had a major impact on the field of medicine since its development more than 40 years ago ^19–21^. Propofol is used for moderate sedation, as well as for induction and maintenance of general anesthesia. Higher doses are sometimes used to induce deep anesthesia for special situations where a decrease in cerebral metabolism is desired (e.g., temporary clipping of a cerebral aneurysm, refractory status epilepticus) ^22,23^. At these higher doses, propofol reliably and safely induces burst suppression, similar to isoflurane.

Many of the original burst-suppression investigations used isoflurane, but propofol treatment is better tolerated. Patients acutely experience pleasant feelings, suffer less nausea, and better maintain blood pressure. In an open-label pilot study of subjects with treatment-resistant depression, we found that a series of brief infusions with propofol, dosed to EEG burst suppression, reduced depression severity by 58% on average, and that depression improved similarly to a comparison group who received ECT ^24^.

Those findings prompted the current study, which more rigorously examined the clinical antidepressant effects of propofol using a controlled, randomized, blinded trial design. Due to practical considerations, this small trial was designed primarily to gather preliminary data on clinical efficacy of two different doses of propofol for depression, and to assess the feasibility and safety of a larger trial. This trial gathered key information related to blinding, recruitment, tolerability, anesthetically active placebos, and cardiovascular and other safety considerations. Participants were randomized to a high-dose propofol intervention that induced burst suppression versus a low-dose propofol intervention that produced moderate sedation without burst suppression. Quantitative real-time EEG was used to guide dosing to the specified pharmacodynamic targets. We hypothesized that subjects randomized to high dose propofol with burst suppression would experience greater improvement in depression than those randomized to the low-dose treatment (moderate sedation without burst suppression), that these treatments could be safely administered without serious adverse events, and that treatments would be well tolerated and acceptable to patients.

## Methods

### Overall study design

This research was approved by the University of Utah Institutional Review Board and preregistered at ClinicalTrials.gov (NCT03684447). Subjects provided written informed consent. The trial used a randomized, blinded, controlled design with two parallel arms: low-dose propofol versus high-dose propofol (1:1 allocation). Each intervention consisted of a series of 6 infusions, delivered 3 times per week. Subjects who were initially randomized to low dose and did not respond after 6 treatments had the option to continue with a series of 6 open-label high-dose treatments. See *Supplemental Methods* for additional design details.

### Participants and baseline assessments

Treatment-seeking adult outpatients with a current moderate-to-severe depressive episode were recruited from the Treatment Resistant Mood Disorder Clinic at the University of Utah. Patients at elevated risk for anesthetic complications or suicide were excluded (see Supplementary Table S1 for complete criteria). The baseline assessment included a preoperative medical evaluation, the Mini International Neuropsychiatric Interview (M.I.N.I. 7.0.0), the 24-item Hamilton Depression Rating Scale (HDRS-24), the Patient Health Questionnaire (PHQ-9), the 7-item Generalized Anxiety Disorder scale (GAD-7), the 19-item Beck Scale for Suicide Ideation (BSSI), the 5-item Work and Social Adjustment Scale (WSAS), and the abbreviated World Health Organization Quality of Life scale (WHOQOL-BREF) ^25–31^. See *Supplemental Methods* for details.

### Randomization and blinding

Participants were randomly assigned 1:1 to the low-or high-dose group. The allocation sequence was created with a random-number generator (http://random.org) using a block size of 6; the blocks were manually shuffled so that no one knew the sequence. Staff involved in recruiting participants were not informed of the randomization scheme. Allocation was concealed in an opaque envelope that was opened by the treatment team when a participant arrived for their first infusion session. The anesthesiologist and research assistant performing each treatment were necessarily informed of group assignment due to safety requirements and the individualized nature of the dosing. All others remained blinded: participants, their companions, clinical raters, and other staff. The team of clinical raters was independent of the rest of the research team; raters were not involved in research design, treatment sessions, or team meetings. To reduce the chance of inadvertently unblinding participants through visual cues, the infusion pump was concealed before subjects entered the treatment room and subjects wore an eye covering before and during drug administration.

After completing the Week 2 assessments, the effectiveness of the blind was assessed by asking participants (n = 21) to rate on a 0–100 scale their best guess about which intervention they received: 0 represented certainty it was low dose, 100 certainty it was high dose, and 50 complete uncertainty. After providing this rating, participants were informed of their group assignment.

### Propofol treatments

Treatments were administered in an interventional suite designed for ECT. American Society of Anesthesiologists (ASA) monitoring standards were followed. Intravenous fluid was coloaded to ensure hemodynamic stability. Participants were preoxygenated and anesthesia induced and maintained with propofol. Ondansetron (4 mg) and lidocaine (30 mg) were administered routinely but other medications were not administered unless indicated (see *Supplemental Methods*). Frontal EEG was continuously assessed using a BIS Monitor (BIS VISTA Monitoring System, Aspect Medical Systems) to enable real-time dosing adjustments based primarily on the continuously calculated *suppression ratio (SR*). The SR represents the proportion of time that brain activity is suppressed, ranging from 0 (none) to 100 (complete suppression). The bispectral index (BIS index) is a proprietary unitless depth-of-anesthesia metric based mainly on gamma-band power ^32^. This number was visible but not used to determine patient dosing.

For the high-dose intervention, propofol was individually dosed to EEG burst suppression as in our previous pilot study ^24^, with modifications. An induction bolus of propofol was administered IV over ∼30 seconds and followed by a continuous infusion. Initial dosing of propofol was guided by a population-based pharmacokinetic model ^33^ in order to rapidly reach and maintain a steady-state effect-site concentration ^34^. Concentrations of ∼5–10 mcg/mL are associated with the burst-suppression state ^35,36^. The propofol dose was adjusted in real-time based on quantitative real-time EEG feedback (the SR value). The pharmacodynamic goal was to achieve a SR of 70–90% for 12–15 minutes. Dose adjustments were made with 30–50 mg boluses of propofol, infusion pauses of 1–2 minutes, and/or changes to the infusion rate. We previously found (in the open label pilot study) that the treatments of responders were characterized by shorter duration and less intensity of burst suppression relative to non-responders ^24^. Thus during the current study we avoided burst suppression lasting over 20 minutes and SR values above 90%.

The low-dose propofol intervention was selected due to its pharmacodynamic effects and the lack of current evidence for antidepressant effects. It was intended to simulate the high dose propofol experience (i.e.. sedation/hypnosis, amnesia, emergence) without inducing burst suppression (a suspected mechanism of treatment effects). In this group, propofol was administered IV as a bolus over 10–30 seconds followed by a continuous infusion for 12–15 minutes. The infusion rate was adjusted individually with the goal of achieving moderate sedation, defined by three criteria: (i) a score of 2 or 3 on the Modified Observer’s Assessment of Alertness/Sedation (MOAAS) Scale ^37^; (ii) SR < 10; and (iii) BIS value of 50–90. At this level of sedation, which is associated with an average steady-state effect-site concentration of 1–2 mcg/mL ^33^, subjects were typically momentarily arousable with physical stimulation (e.g., trapezius squeeze).

### Outcome assessments

The primary goal of the study was to assess the feasibility, safety, and preliminary efficacy of propofol anesthesia treatment for patients with treatment resistant depression. The pre-specified primary depression outcome measure was change in total score on the HDRS-24 from baseline to Week 2 (after 6 treatments). The secondary depression outcome was change in PHQ-9 total score at Week 2. The other secondary scales measured anxiety (GAD-7), suicidal ideation (BSSI), functional impairment (WSAS), and quality of life (WHOQOL-BREF). The shorter 17-item and 6-item HDRS scales (HDRS-17 and HDRS-6) were examined in exploratory analyses. Hemodynamic and other physiological and anesthetic treatment related information was collected and analyzed to assess for safety and practicality. See *Supplemental Methods* for additional details.

### Offline EEG analysis

During each treatment session, two channels of left frontal raw EEG data were recorded to flash memory from the BIS Monitor (sampling rate 128 Hz per channel) and data from the first channel were analyzed offline. Each recording was visually inspected and obvious artifacts such as those from BIS Monitor ground checks or patient movement were manually removed. The signal was Butterworth filtered between 0.1 and 45 Hz. A continuous wavelet transform (“cwt”, Wavelet Toolbox, MATLAB, version 2020b) was applied to characterize changes in spectral power over time. Details are reported elsewhere ^38^.

### Statistical analysis

Analyses were performed with R statistical software (version 4.1.2). Prior to study initiation, power simulations indicated that a trial with 12 participants per group was adequately powered to detect a standardized mean difference (SMD) of 1.0 between the low- and high-dose groups (see *Supplemental Methods*). Therefore, we did not anticipate this sample size would provide definitive statistical confirmation of small-or moderate-sized differences between groups. The open-label study results ^24^ suggested that a large effect size was possible for the high-dose group, but the effect size for the low-dose group was unknown. Ultimately the choice of 12 participants per group was determined by practical considerations - i.e., limited resources, time, and funding.

SMD values and confidence intervals were calculated using the “cohen.d” function with Hedges’ correction (“effsize” package, version 0.8.1). The pre-specified primary analysis used a general linear model (“lm” function, base R) to test the hypothesis that depression severity would improve more in the high-dose group than in the low-dose group after 6 treatments. HDRS-24 total score at Week 2 was modeled as the response variable, group assignment as the predictor of interest, and baseline HDRS-24 score as a covariate. A main effect of group at p < 0.05 (two-sided) was considered evidence in support of the primary hypothesis. The same kind of linear model was used to test the secondary depression outcome measure, change in PHQ-9 total score. See *Supplemental Methods* for additional details.

## Results

### Participants

Twenty-four subjects were randomized and all completed the series of 6 treatments and the primary Week 2 outcome assessments (see Supplemental Figure S1). Low- and high-dose groups were similar with respect to baseline demographic and clinical characteristics (Table 1).

**Table 1.** Demographic and baseline clinical characteristics of participants.

|  | Low Dose (n=12) |  | High Dose (n=12) |  | P |
| --- | --- | --- | --- | --- | --- |
| Demographics |  |  |  |  |  |
| Age, years | 41.5 | (10.6) | 34.7 | (7.5) | 0.09 |
| Sex assigned at birth, female, n (%) | 5 | (41.7) | 6 | (50.0) | 1.00 |
| Current gender, female, n (%) | 4 | (33.3) | 5 | (41.7) | 1.00 |
| Self-reported race, white, n (%) | 11 | (91.7) | 12 | (100.0) | 1.00 |
| Married or domestic partner, n (%) | 5 | (41.7) | 4 | (33.3) | 1.00 |
| Education, years | 16.9 | (2.1) | 15.8 | (2.5) | 0.23 |
| Annual household income level <sup>a</sup> | 4.0 | (1.8) | 4.3 | (2.1) | 0.73 <sup>b</sup> |
| Illness severity |  |  |  |  |  |
| 24-item Hamilton Depression Rating Scale (HDRS-24) | 28.3 | (5.7) | 30.9 | (7.6) | 0.36 |
| 17-item Hamilton Depression Rating Scale (HDRS-17) | 19.2 | (4.5) | 21.8 | (6.3) | 0.25 |
| 6-item Hamilton Depression Rating Scale (HDRS-6) | 10.6 | (1.8) | 11.5 | (2.4) | 0.30 |
| Quick Inventory of Depressive Symptomatology (QIDS-SR) | 18.7 | (4.1) | 18.3 | (3.3) | 0.79 |
| Patient Health Questionnaire (PHQ-9) <sup>c</sup> | 19.9 | (3.4) | 19.4 | (4.0) | 0.75 |
| Work and Social Adjustment Scale (WSAS) <sup>c</sup> | 32.1 | (3.6) | 31.4 | (5.2) | 0.72 |
| Global Assessment of Functioning scale (GAF) | 50.0 | (5.8) | 50.6 | (6.0) | 0.81 |
| Clinical Global Impression Severity, 4 moderately ill, n (%) | 2 | (16.7) | 2 | (16.7) | 1.00 |
| 5 markedly ill, n (%) | 6 | (50.0) | 6 | (50.0) |  |
| 6 severely ill, n (%) | 4 | (33.3) | 4 | (33.3) |  |
| Generalized Anxiety Disorder scale (GAD-7) <sup>c</sup> | 10.7 | (5.5) | 12.2 | (4.1) | 0.49 |
| Beck Scale for Suicide Ideation (BSSI) <sup>c</sup> | 7.3 | (9.0) | 9.6 | (8.1) | 0.53 |
| WHO Quality of Life, Physical domain <sup>c</sup> | 40.9 | (12.0) | 44.1 | (15.6) | 0.59 |
| WHO Quality of Life, Psychological domain <sup>c</sup> | 20.8 | (12.1) | 22.6 | (13.5) | 0.75 |
| WHO Quality of Life, Social domain <sup>c</sup> | 35.6 | (17.5) | 38.2 | (21.5) | 0.75 |
| WHO Quality of Life, Environmental domain <sup>c</sup> | 59.7 | (16.9) | 62.2 | (17.5) | 0.72 |
| Chronicity and resistance |  |  |  |  |  |
| Depressive episode duration, months | 19.9 | (14.5) | 10.6 | (8.5) | 0.13 <sup>b</sup> |
| Chronic episode (>2 years), n (%) | 4 | (33.3) | 2 | (16.7) | 0.64 |
| Age of depression onset, years | 16.9 | (4.2) | 15.9 | (7.3) | 0.68 |
| Maudsley stage | 7.8 | (1.4) | 7.7 | (1.7) | 0.80 |
| Massachusetts General Hospital stage | 3.8 | (1.7) | 4.4 | (1.7) | 0.41 |
| History of electroconvulsive therapy, n (%) | 2 | (16.7) | 2 | (16.7) | 1.00 |
| Other clinical features |  |  |  |  |  |
| Bipolar disorder, n (%) | 1 | (8.3) | 1 | (8.3) | 1.00 |
| Generalized anxiety disorder, n (%) | 7 | (58.3) | 6 | (50.0) | 1.00 |
| Panic disorder, n (%) | 1 | (8.3) | 1 | (8.3) | 1.00 |
| Agoraphobia, n (%) | 3 | (25.0) | 2 | (16.7) | 1.00 |
| Social anxiety disorder, n (%) | 2 | (16.7) | 6 | (50.0) | 0.19 |
| Eating disorder, n (%) | 2 | (16.7) | 2 | (16.7) | 1.00 |
| Lifetime history of psychosis, n (%) | 1 | (8.3) | 0 | (0.0) | 1.00 |
| Maternal mood disorder, n (%) | 6 | (50.0) | 9 | (75.0) | 0.40 |
| Paternal mood disorder, n (%) | 2 | (16.7) | 4 | (33.3) | 0.64 |
| Body mass index, kg/m<sup>2</sup> | 30.5 | (6.7) | 27.1 | (5.6) | 0.19 |

P values derive from two-sample t tests for continuous variables or Fisher exact tests for categorical variables, except where indicated. Other values indicate mean (SD) except where indicated. <sup>a</sup> Income level was an ordinal variable ranging 1-7 (<\$20,000 to >\$120,000). <sup>b</sup> Wilcoxon rank sum test. <sup>c</sup> Data missing for one low-dose subject. WHO, World Health Organization.

### Treatment characteristics

The low-dose propofol intervention produced moderate-to-deep sedation without EEG burst suppression, as intended (Figure 1). As shown in Table 2, the mean value for the BIS index was 59.3. MOAAS scores averaged 1.6, which was lower than the anticipated target of 2–3. Nonetheless, these values were consistent with moderate sedation, and no burst suppression was observed during any low-dose infusion.

**Figure 1.**
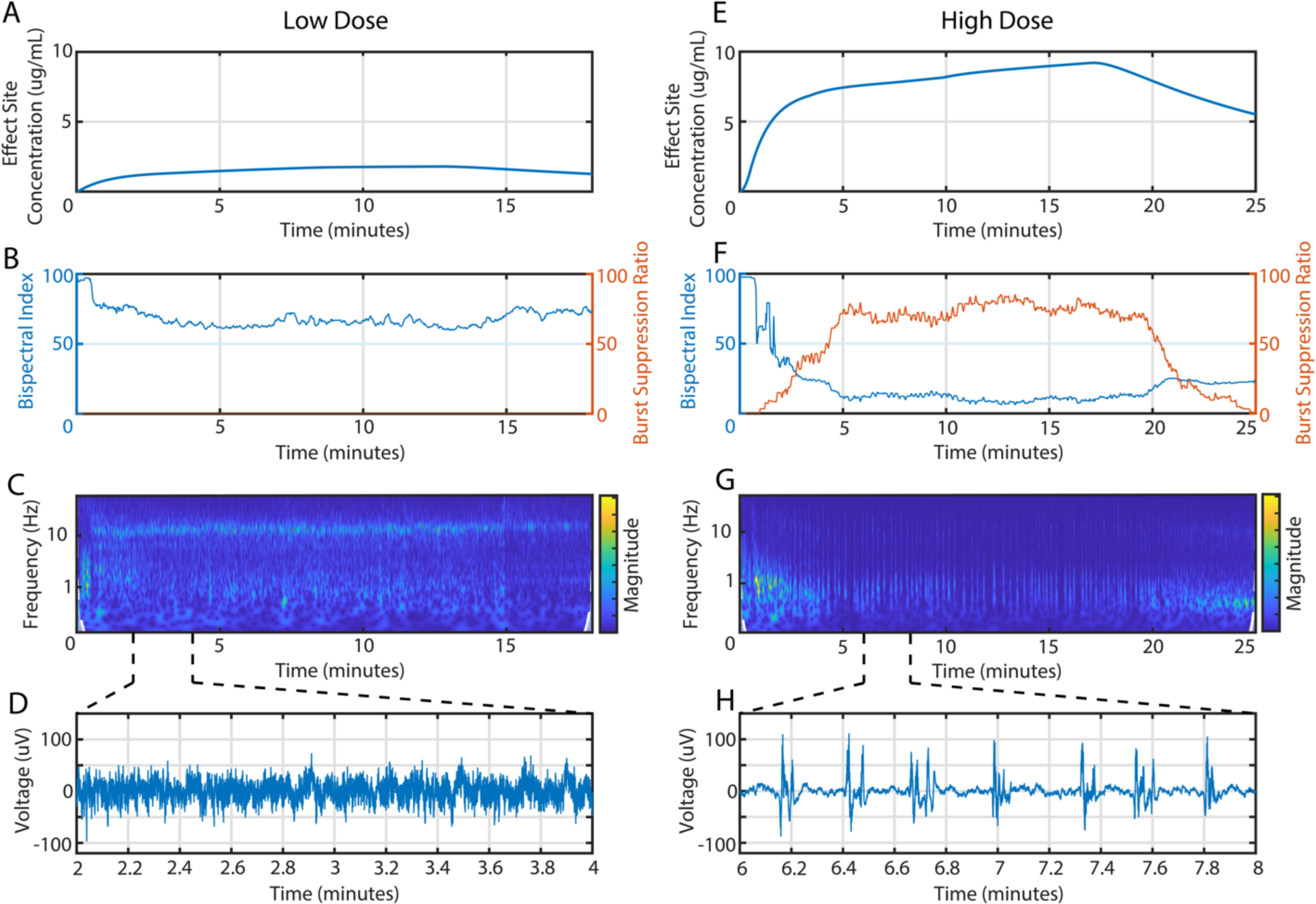
Real-time electroencephalography (EEG) guided propofol treatments. ***A-D***, example of a Low-Dose treatment session. ***E-H***, example of a High-Dose treatment session. (***A***, ***E***) Estimated effect site concentration based on actual dosing and a population-based pharmacokinetic model. (***B***, ***F***) BIS index and SR index calculated in real-time based on frontal EEG. (***C***, ***G***) Spectrogram calculated with a continuous wavelet transform showing frequency content of the signal over time. (***D***, ***H***) Excerpts of EEG signals contrasting moderate sedation versus burst suppression.

**Table 2.** Characteristics of treatment sessions.

|  | Low Dose (n=12) |  | High Dose (n=12) |  | P |
| --- | --- | --- | --- | --- | --- |
| **Propofol dosing** |  |  |  |  |  |
| Induction dose, mg | 77 | (17) | 335 | (56) | <0.0001 |
| Sum of bolus doses, mg | 78 | (18) | 366 | (67) | <0.0001 |
| Dose delivered via infusion, mg | 106 | (35) | 405 | (98) | <0.0001 |
| Total dose administered, mg | 183 | (49) | 772 | (156) | <0.0001 |
| Total dose administered, mg/kg | 1.98 | (0.40) | 9.30 | (1.60) | <0.0001 |
| Estimated effect site concentration, mean, ug/ml | 1.83 | (0.36) | 6.62 | (0.78) | <0.0001 |
| Estimated effect site concentration, peak, ug/ml | 1.97 | (0.40) | 8.24 | (1.04) | <0.0001 |
| **Mean arterial pressure** |  |  |  |  |  |
| Pre-treatment baseline, mmHg | 92.9 | (12.7) | 94.1 | (8.9) | 0.80 |
| Intraoperative minimum, mmHg | 78.4 | (8.4) | 67.9 | (5.8) | 0.002 |
| Intraoperative minimum relative to baseline | 0.848 | (0.035) | 0.727 | (0.050) | <0.0001 |
| **Pharmacodynamic parameters** |  |  |  |  |  |
| Cumulative EEG suppression (SR integral), minutes | 0 | (0) | 12.23 | (1.47) | - |
| Duration of EEG burst-suppression state, minutes | - | - | 17.03 | (1.32) | - |
| Mean intensity of EEG burst-suppression state (SR) | - | - | 68.3 | (6.2) | - |
| Time spent with SR 70–90, minutes | - | - | 8.06 | (2.97) | - |
| Time spent with SR 50–70, minutes | - | - | 5.52 | (2.26) | - |
| Time spent with SR 50–90, minutes | - | - | 13.58 | (1.95) | - |
| Time spent with SR over 90, minutes | - | - | 0.93 | (0.81) | - |
| Mean BIS index | 59.3 | (5.2) | - | - | - |
| Mean MOAAS score | 1.59 | (0.75) | - | - | - |
| **Procedure durations** |  |  |  |  |  |
| Propofol infusion, minutes | 12.13 | (0.23) | 15.04 | (0.89) | <0.0001 |
| End of infusion to emergence, minutes | 7.93 | (2.52) | 26.50 | (4.00) | <0.0001 |
| Start of infusion to emergence, minutes | 19.41 | (2.46) | 40.66 | (4.76) | <0.0001 |

Characteristics were calculated per treatment session, and then averaged across 6 sessions for each subject. Values indicate mean (SD) across subjects. P values represent two-sample t tests. Cumulative EEG suppression was the integral of the SR curve across the entire session. The burst-suppression state was defined as the period of time during which SR > 30.

High-dose propofol treatments elicited deep general anesthesia with EEG burst suppression (Figure 1). The average time spent with a suppression ratio (SR) of 70–90 was 8 minutes per treatment session, which was lower than the intended 12–15 minutes (Table 2). However, the SR dwelled in the broader range of 50–90 for an average of 14 minutes, which was consistent with the targeted duration (Table 2). For some participants the SR parameter computed in real time by the BIS Monitor appeared to underestimate the degree of suppression (further described elsewhere ^38^) so in these cases SR of 50–70% was visually targeted to avoid excessive cortical suppression.

### Primary depression outcome

#### Low-dose group

The mean [95%-CI] change in HDRS-24 total score from baseline to Week 2 was −5.3 [−10.3, −0.2] points (p = 0.04, t = 2.29, df = 11, paired t-test). This change corresponded to a 17% decrease in depression severity and an effect size of −0.77 [−1.56, 0.24] (Figure 2A, Figure 3, Supplementary Figure S2, Supplementary Table S2).

**Figure 2.**
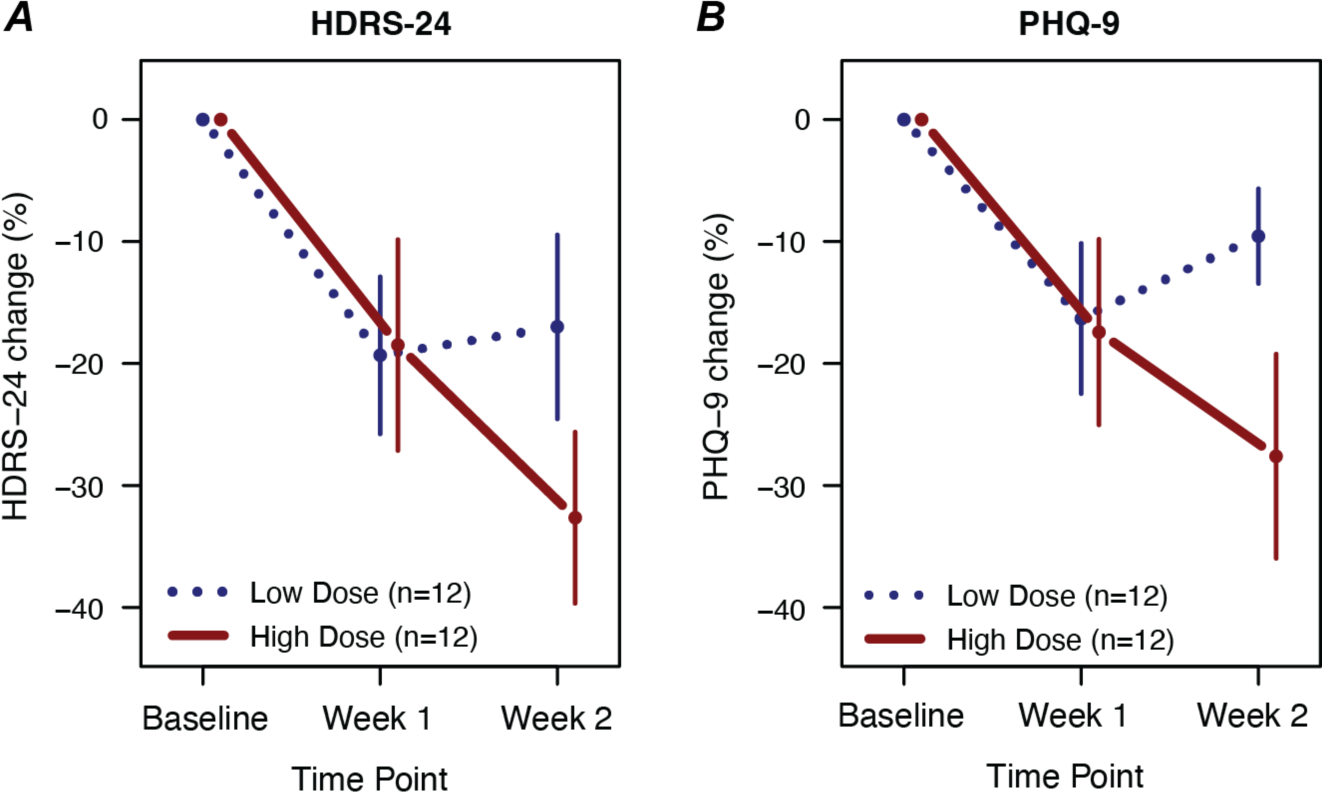
Change in depression severity with Low-Dose versus High-Dose propofol treatment. (***A***) Change in 24-item Hamilton Depression Rating Scale (HDRS-24) score relative to Baseline at Week 1 (after 3 treatments) and Week 2 (after 6 treatments) for the two treatment groups. (***B***) Change in Patient Health Questionnaire (PHQ-9) score. Mean +/-SEM shown. Group differences at Week 2 did not reach statistical significance (p > 0.05).

**Figure 3.**
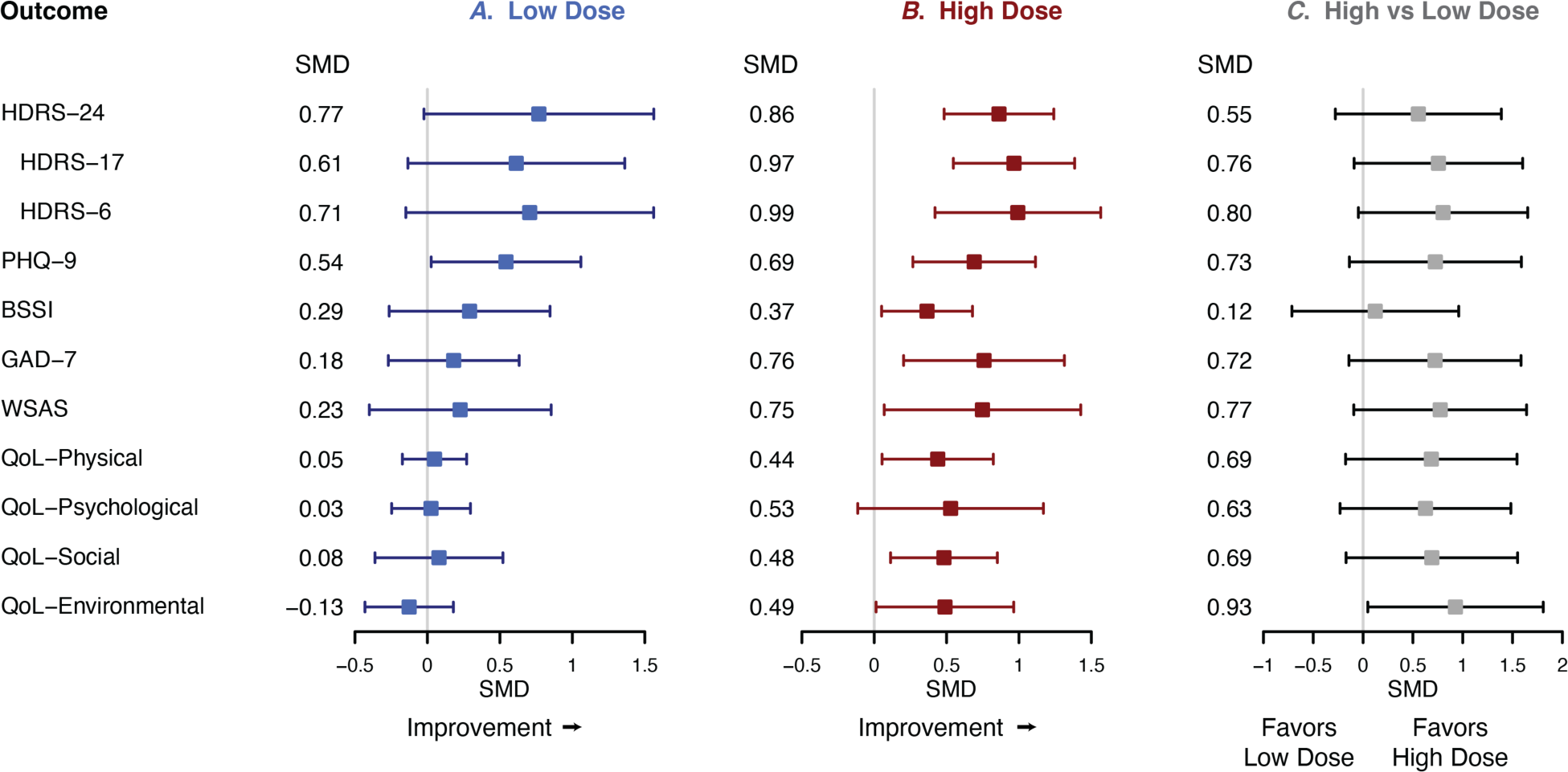
Effect sizes for changes in clinical outcomes. Standardized mean differences (SMD) and 95% confidence intervals are shown for (***A***) the Low-Dose group (n=12), (***B***) the High-Dose group (n=12), and (***C***) the contrast of High Dose versus Low Dose. (***A***, ***B***) Paired SMD values for the Low-Dose and High-Dose groups indicate change from baseline to Week 2. For each scale, improvement is represented by positive values. (***C***) SMD values for High versus Low Dose indicate between-group differences in change scores. For each scale, positive values indicate greater improvement among the High-Dose group. HDRS, Hamilton Depression Rating Scale (24-, 17-, and 6-item versions). PHQ-9, Patient Health Questionnaire. BSSI, Beck Scale for Suicide Ideation. GAD-7, Generalized Anxiety Disorder scale. WSAS, Work and Social Adjustment Scale. QoL, abbreviated World Health Organization Quality of Life scale (WHOQOL-BREF).

#### High-dose group

The mean change in HDRS-24 score was −9.3 points [−12.9, −5.6] (p = 0.0002, t = 5.52, df = 11, paired t-test). This change corresponded to 33% improvement and an effect size of −0.86 [−1.24, −0.48] (Figure 2A, Figure 3, Supplementary Figure S2, Supplementary Table S2).

#### Group comparison

Change in HDRS-24 score from baseline to Week 2 did not show statistical significance between groups (β = −3.51, SEM = 2.91, p = 0.24, general linear model). The between-group effect size was −0.55 [−1.39, 0.28] (Supplementary Table S2, Figure 3). Similarly, a linear mixed model including HDRS-24 score at baseline, Week 1, and Week 2 revealed no significant group-by-time interaction (p = 0.14, chi-squared = 2.14, df = 1).

#### Exploratory analyses

The abbreviated HDRS-17 and HDRS-6 scales showed that, compared to the HDRS-24, between-group effect sizes for those measures tended to be greater in magnitude (−0.76 and −0.80, respectively; Supplementary Table S2).

### Secondary depression outcome

#### Low-dose group

The mean [95%-CI] change in PHQ-9 score was −2.0 points [−3.9, −0.1] (p = 0.04, t = 2.35, df = 10, paired t-test, 1 missing value). That corresponded to a −10% change and an effect size of −0.54 (Figure 2B, Supplementary Table S2, Figure 3).

#### High-dose group

The mean change in PHQ-9 score was −4.8 points [−7.7, −2.0] (p = 0.003, t = 3.76, df = 11, paired t-test), which corresponded to a −28% change and an effect size of −0.69 (Figure 2B, Supplementary Table S2, Figure 3).

#### Group comparison

Change in PHQ-9 showed a non-significant trend favoring the high-dose group (β = −2.81, SEM = 1.61, p = 0.096, general linear model). The effect size [95%-CI] between groups was −0.73 [−1.59, 0.14] (Supplementary Table S2, Figure 3). Similarly, a linear mixed model including PHQ-9 score at baseline, Week 1, and Week 2 revealed a trend-level group-by-time interaction (p = 0.074, chi-squared = 3.19, df = 1).

### Outcomes beyond depression

Other secondary outcomes included measures of suicidal ideation, anxiety, functional impairment, and quality of life (Supplementary Table S2, Figure 3). Changes in BSSI score from baseline to Week 2 were similar for low-dose versus high-dose groups (p = 0.77, Supplementary Table S2). Changes in GAD-7, WSAS, and WHOQOL-BREF domains favored the high-dose condition with effect-size magnitudes > 0.6, but only the WHOQOL-BREF environmental domain reached statistical significance (p = 0.03, Supplementary Table S2).

### Adverse effects

The most common adverse effects reported during recovery were sore throat, grogginess, fatigue, and discomfort at the IV catheter site (Supplementary Table S3). Sore throat, delayed reorientation, and moderate (but well tolerated) blood pressure decrease were more likely in the high-dose group. All side effects were mild and/or self-limited (Supplementary Table S3, Table 2).

### Effectiveness of blinding

Following the Week 2 assessment, participants were asked to guess which intervention they received on a 0–100 scale, 50 representing complete uncertainty. The mean (SD) rating was 42 (18) for the low-dose group and 53 (14) for the high-dose group; neither was significantly different from 50 (p = 0.16 and p = 0.52, one-sample t-tests). Similarly, neither HDRS-24 improvement nor PHQ-9 improvement were associated with guesses (p > 0.15, linear regression). We conclude that blinding procedures during the trial were successful.

### Open-label high-dose treatment outcomes

Ten of the 12 participants who initially received low-dose propofol during the blinded phase of the trial were eligible to continue with a series of open-label high-dose treatments, and 7 subjects completed the 6 treatments per protocol (Supplementary Figure S1). The mean [95%-CI] change in HDRS-24 score from Week 2 to Week 4 time points was −12.5 points [−19.2, −5.8], corresponding to a 49% improvement (p = 0.0095, t = 5.95, df = 3, paired t-test, 3 missing values). The mean [95%-CI] change in PHQ-9 score was −7.5 points [−12.8, −2.2], representing a decrease of 40% (p = 0.015, t = 3.61, df = 5, paired t-test, 1 missing value).

## Discussion

To our knowledge, this study is the first controlled trial to evaluate the clinical antidepressant effects of propofol. We utilized real-time EEG to deliver propofol to meet predefined pharmacodynamic criteria. This approach allowed us to accommodate inter-subject variability in pharmacokinetics and pharmacodynamics, and to confirm that we engaged the brain target as intended. Both low- and high-dose infusions were well tolerated, and the protocols used were effective in maintaining the blind. Depression, anxiety, functional impairment, and quality of life improved in both dosing groups after a series of 6 infusions and, as hypothesized, the degree of improvement was nominally greater for the high-dose group.

The group difference was not statistically significant, which is unsurprising given the low power of this study to detect small-to-medium sized effects. Nonetheless, these results demonstrate the feasibility of a rigorous controlled trial of propofol, as well as the effectiveness of low dose administration as a pharmacologically but not psychiatrically active “placebo” and they provide preliminary support for the notion that propofol may have dose-dependent antidepressant effects. The findings lay the groundwork for a larger trial to definitively evaluate the therapeutic efficacy of propofol. The tolerability and low rate of side effects that we observed suggest propofol could represent a viable alternative to ECT that lacks ECT-related cognitive adverse effects.

It is instructive to contrast the current controlled trial to our previous open-label trial of propofol ^24^. Here we administered a series of 6 infusions rather than 10 in the original study, due to concerns about feasibility and acceptability. Specifically, we decided against a design in which participants might complete 10 treatments and then have to cross over to the other dose for 10 more treatments, due to subject burden. HDRS-24 scores improved by 33% on average among subjects randomized to 6 high-dose infusions, which was less than the 48% improvement observed after 5 treatments in the previous trial. A possible explanation for this difference is that the experimental design of the current trial reduced the magnitude of the placebo effect (intrinsic to all depression trials) by decreasing positive expectancy.

Additionally, the treatments delivered in the current study were less intense (less time spent in the suppressed state) than in the original trial. This design decision was based on the post hoc observation that less intense treatment sessions were associated with better antidepressant response and less hypotension in the original trial ^24^; the goal was to minimize any toxic effects due to excessive and unnecessary dosing. The mean SR intensity was 68 in the current study versus 90 in the previous trial (range 53–78 versus 81–97). It could be that the lower intensities of burst suppression used in the current trial have weaker antidepressant effects. Consistent with this idea, the trajectory of depression severity in the high-dose group, but not the low-dose group, was downward after 6 treatments (see Figure 2), suggesting that additional treatments may have produced further improvement. Taken together, these observations suggest that at least 9 treatment sessions with an SR intensity of 80–90 for 15 minutes might be an optimal high-dose intervention.

The low-dose intervention designed for this trial proved to be a suitable comparison condition. We considered other active comparators such as intravenous midazolam, but the rapid offset of action of propofol is not easily simulated with other drugs. We expected that a moderate dose of propofol would be unlikely to have strong antidepressant effects and would resemble high-dose propofol sufficiently to maintain blinding. Those expectations were borne out. The mean improvement in depression severity was < 20% in the low-dose group.

Participants in the low-dose group were moderately sedated and all had the experience of falling asleep and then waking up. Subjects were unable to accurately guess whether they had received the low-dose or high-dose intervention during the randomized phase. It is important to note, however, that subjects who were exposed to *both* types of treatment were able to distinguish them based on the recovery experience, so a cross-over trial of different propofol doses would likely be compromised by unblinding at the point of cross-over.

Secondary outcomes included self-rated measures of depression (PHQ-9), suicidal ideation (BSSI), anxiety (GAD-7), functional impairment (WSAS), and quality of life (WHOQOL-BREF) as well as the two commonly used abbreviated versions of the HDRS. The smallest group difference (effect size −0.12) was seen for the BSSI. All other secondary measures favored the high-dose group, with estimated effect-size magnitudes of 0.63–0.93 (albeit with wide confidence intervals). These effect sizes are comparable to those observed with a single ketamine infusion ^39^. Among the largest group differences (effect size −0.80) was that for the HDRS-6, which emphasizes core depressive symptoms; the difference between groups was marginally significant (p = 0.05). Collectively this pattern of findings suggests that high-dose propofol has the greatest effects on core depressive symptoms, but that broader effects on anxiety and functioning are also likely. The results also suggest that high dose propofol is unlikely to have specific anti-suicide effects beyond the non-specific effects due to participating in a clinical trial.

A significant limitation of this trial was the limited sample size (12 per group) which was adequately powered to detect only large differences between high-and low-dose groups (standardized mean difference of 1.0 or greater). Similarly, the series of 6 treatments may have been sub-optimal for eliciting antidepressant effects. These limitations were largely due to the intensive nature of the interventions and limited resources: anesthesiologists were required to deliver nearly 200 infusions during this trial. An adequately powered study would likely require more than double that number of treatments. Other notable limitations are the enrollment of participants at a single academic center, the lack of racial and ethnic diversity in the sample, and the need to exclude participants with significant medical comorbidities, which may limit the generalizability of the findings.

In conclusion, this study demonstrated the feasibility, tolerability, and safety of EEG-guided propofol infusion and its potential as a therapeutic intervention for treatment resistant depression. Although these findings suggest clinically significant antidepressant effects of high dose propofol infusions, a larger clinical trial will be needed to more definitively answer this question. Subsequent studies are warranted to further characterize how a burst-suppression state may elicit therapeutic neuroplastic and antidepressant effects. This work is the first controlled trial of propofol for depression and it provides additional evidence for the introduction of a new age of investigation to explore “therapeutic anesthesia” - the potential for psychoactive and consciousness-altering anesthetic medications to therapeutically induce cerebral plasticity, resulting in long lasting improvements in mood and quality of life.

## Data Availability

All data produced in the present study are available upon reasonable request to the authors

## Acknowledgements

We thank the patients, families, and staff of the Treatment Resistant Mood Disorder clinic for their enthusiastic support of this work. We acknowledge valuable advice, support, and guidance from Lowry Bushnell, Alan Light, Talmage Egan, and Nathan Pace. We dedicate this work to the memory of Michael Cahalan.

---

**Supplementary Figure S1.**
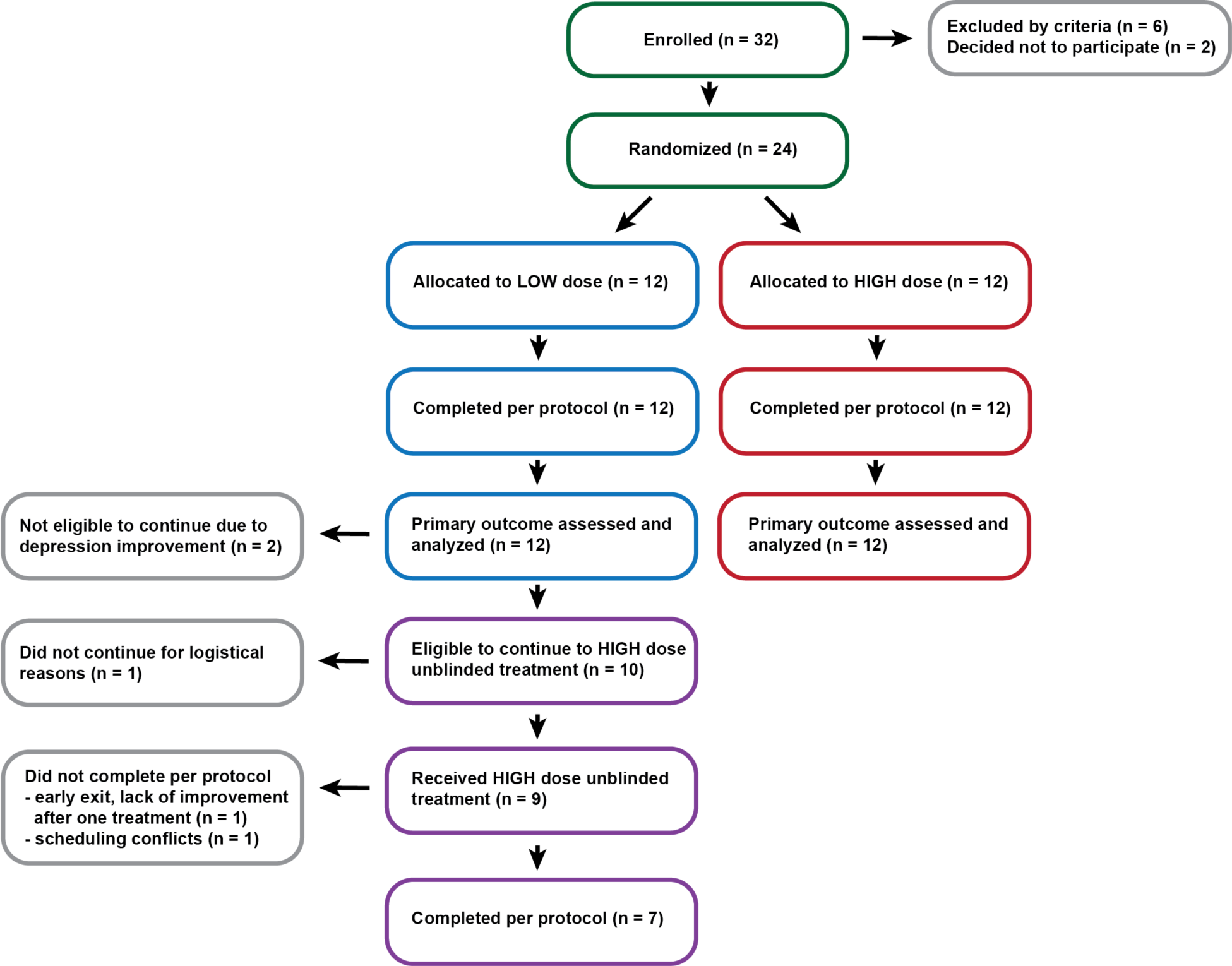
Participant flow diagram.

**Supplementary Figure S2.**
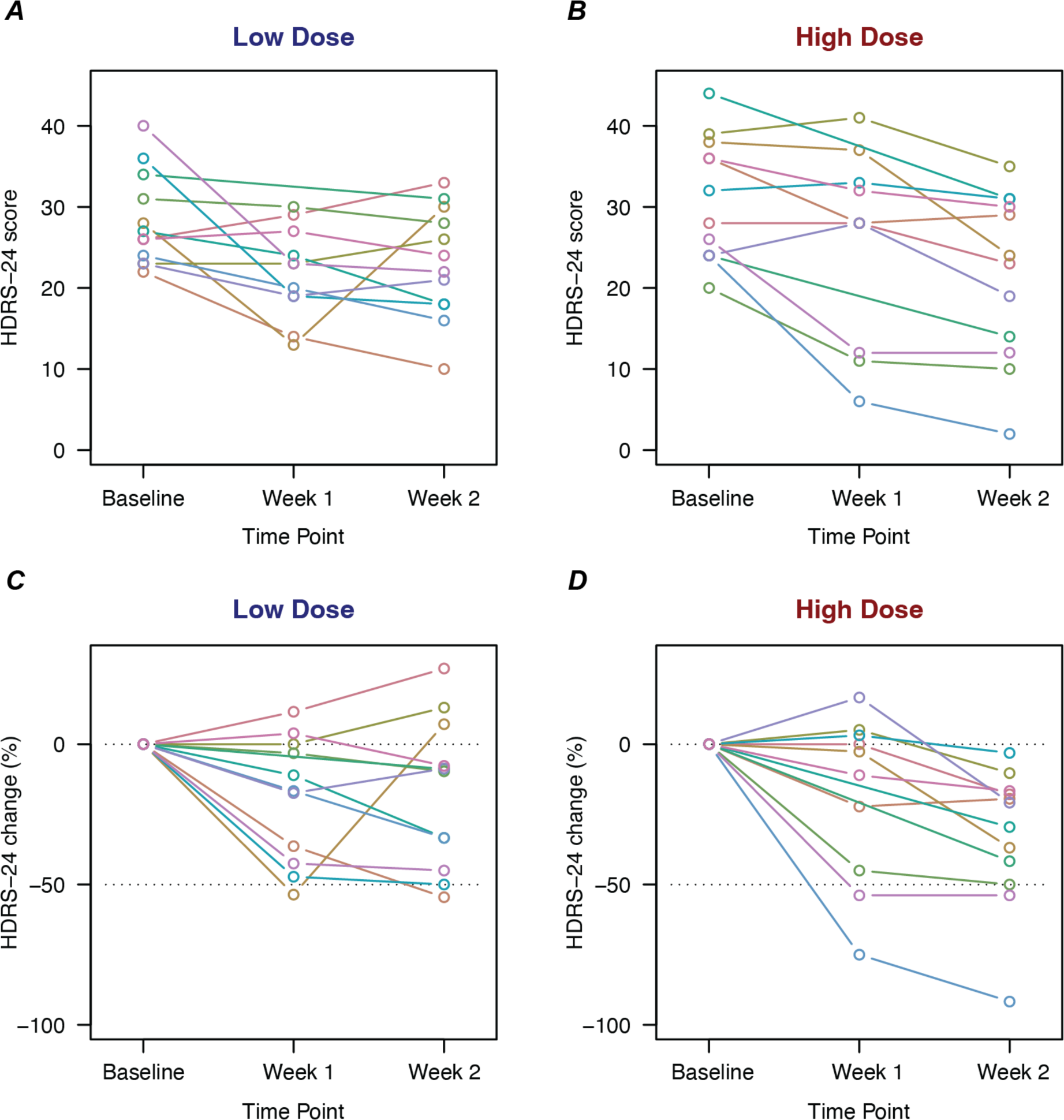
Hamilton Depression Rating Scale scores for individual participants. (***A***) Scores for subjects in the Low-Dose group are shown at Baseline, Week 1 (after 3 treatments), and Week 2 (after 6 treatments). (***B***) Scores for subjects in the High-Dose group. (***C***) Scores for the Low-Dose group expressed as change from Baseline. (***D***) Change from Baseline for the High-Dose group.

**Supplementary Table S1.**
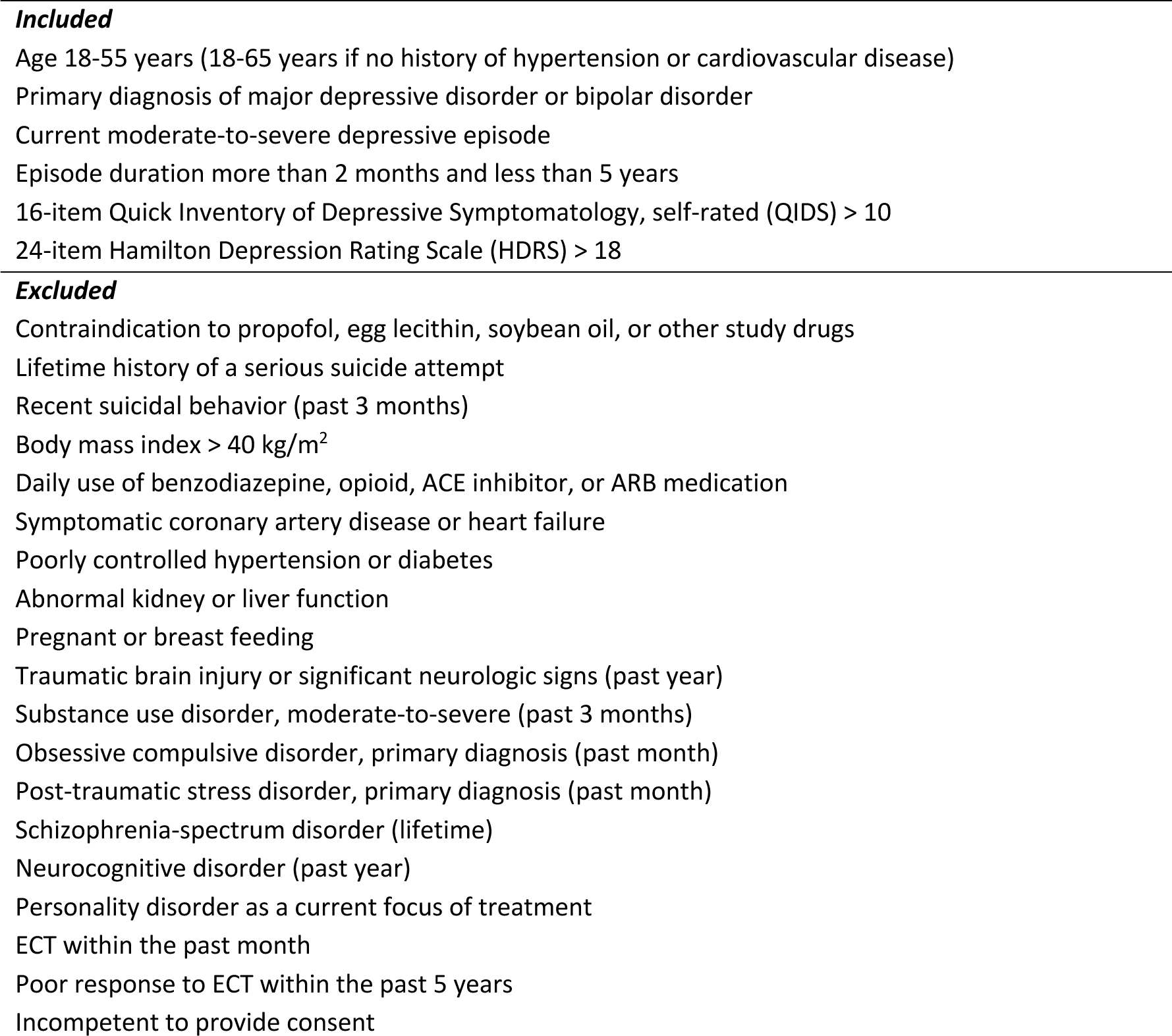
Participant inclusion and exclusion criteria.

**Supplementary Table S2.** Changes in clinical outcome measures from Baseline to Week 2.

| Outcome measure | Low Dose (n = 12) |  |  | High Dose (n = 12) |  |  | High vs Low Dose |  |  |  |  |  |
| --- | --- | --- | --- | --- | --- | --- | --- | --- | --- | --- | --- | --- |
|  | Change | 95%-CI |  | Change | 95%-CI |  | SMD | 95%-CI |  | t | df | P |
| HDRS-24 | -5.25 | -10.30 | -0.20 | -9.25 | -12.94 | -5.56 | -0.56 | -1.39 | 0.28 | -1.41 | 20.1 | 0.174 |
| HDRS-17 | -3.25 | -7.11 | 0.61 | -7.42 | -10.24 | -4.60 | -0.76 | -1.60 | 0.09 | -1.92 | 20.1 | 0.069 |
| HDRS-6 | -1.50 | -3.22 | 0.22 | -3.92 | -5.88 | -1.95 | -0.80 | -1.65 | 0.05 | -2.04 | 21.6 | 0.054 |
| PHQ-9 | -2.00 | -3.90 | -0.10 | -4.83 | -7.66 | -2.01 | -0.73 | -1.59 | 0.14 | -1.84 | 18.8 | 0.082 |
| BSSI | -2.55 | -7.63 | 2.54 | -3.33 | -6.28 | -0.39 | -0.12 | -0.96 | 0.72 | -0.30 | 16.3 | 0.770 |
| GAD-7 | -1.00 | -3.64 | 1.64 | -4.17 | -7.02 | -1.31 | -0.72 | -1.59 | 0.14 | -1.81 | 21.0 | 0.086 |
| WSAS | -1.09 | -4.28 | 2.10 | -7.50 | -13.89 | -1.11 | -0.77 | -1.64 | 0.09 | -1.98 | 16.0 | 0.065 |
| WHOQOL-BREF, Physical | 0.97 | -3.71 | 5.66 | 7.14 | 0.80 | 13.49 | 0.69 | -0.18 | 1.55 | 1.73 | 19.7 | 0.100 |
| WHOQOL-BREF, Psychological | 0.38 | -4.04 | 4.80 | 8.68 | -1.82 | 19.18 | 0.63 | -0.23 | 1.48 | 1.61 | 14.7 | 0.129 |
| WHOQOL-BREF, Social | 1.52 | -7.45 | 10.48 | 11.11 | 2.56 | 19.66 | 0.69 | -0.17 | 1.55 | 1.72 | 20.9 | 0.101 |
| WHOQOL-BREF, Environmental | -2.27 | -8.14 | 3.60 | 8.59 | 0.21 | 16.98 | 0.93 | 0.05 | 1.81 | 2.35 | 19.2 | 0.030 |

Change scores and 95% confidence intervals (95%-CI) were calculated with paired t tests. Standardized mean differences (SMD) and confidence intervals were calculated using Hedges' correction. Welch two-sample t tests (t, df, P values) were applied to compare change scores between high-dose and low-dose groups. HDRS, Hamilton Depression Rating Scale (24-, 17-, and 6-item versions). PHQ-9, Patient Health Questionnaire. BSSI, Beck Scale for Suicide Ideation. GAD-7, Generalized Anxiety Disorder scale. WSAS, Work and Social Adjustment Scale. WHOQOL-BREF, abbreviated World Health Organization Quality of Life scale.

**Supplementary Table S3.**
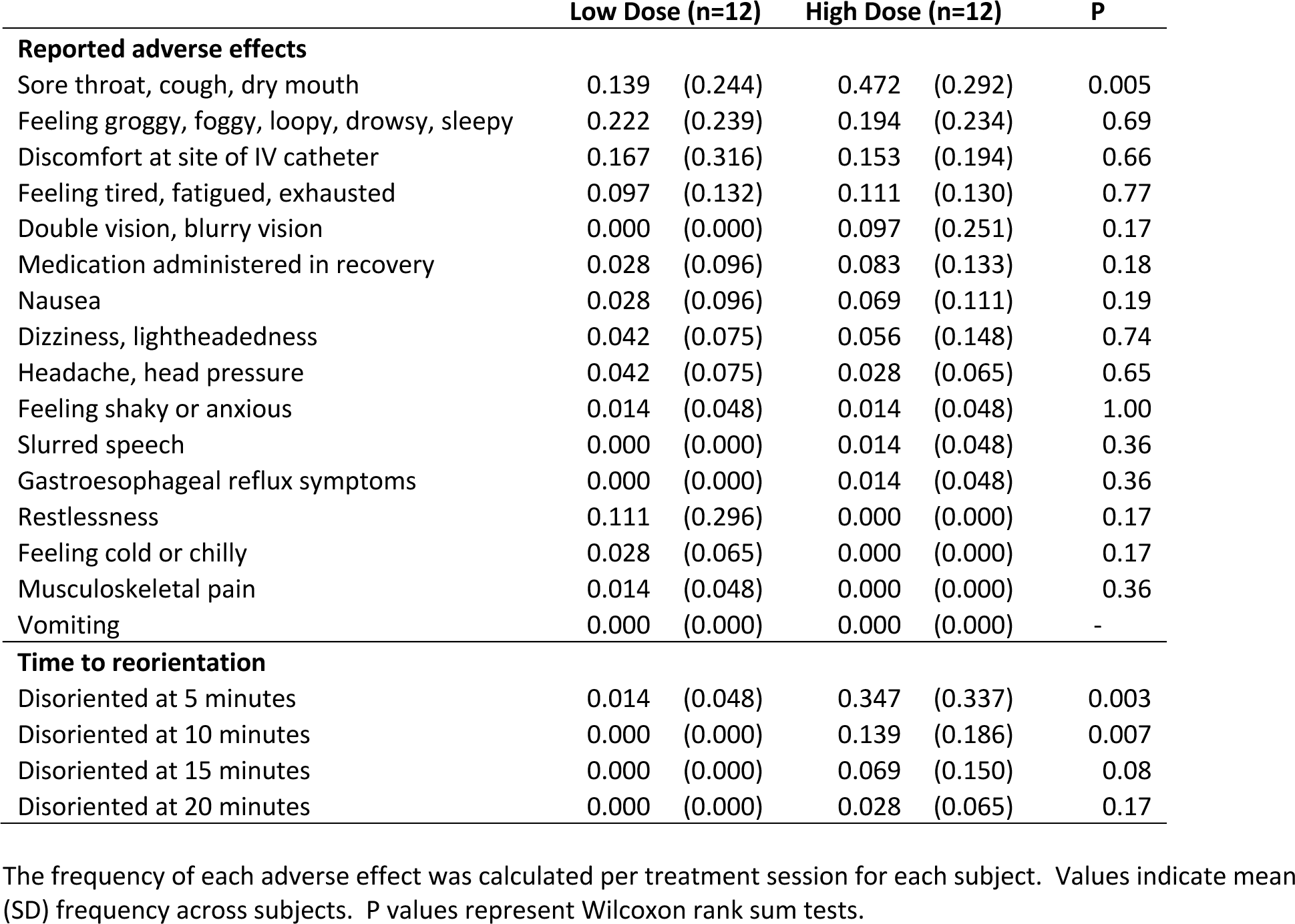
Adverse effects during the recovery phase.

