## Supplemental Methods for "Propofol for treatment resistant depression: A randomized controlled trial"

Supplemental Material

### Supplemental Methods

#### *Study design*

The Neural and Antidepressant Effects of Propofol (NAP) study was conducted between January 2020 and September 2022. The study had two major aims: to evaluate the clinical antidepressant effects of EEG-guided propofol infusion (high-dose versus low-dose) and to characterize changes in associated biomarkers. Here we report findings related to the first aim. An independent three-member Data and Safety Monitoring Board formally monitored study progress and adverse events.

The trial employed a randomized, blinded, controlled design with two parallel arms. Participants were allocated 1:1 to a low-dose propofol intervention versus a high-dose propofol intervention. We decided against comparing propofol to a different active comparator drug (e.g., midazolam sedation) because the antidepressant effects of low-dose propofol were of intrinsic interest and because blinding was facilitated by using propofol in both arms of the trial. Both interventions consisted of a series of 6 infusions, delivered 3 times per week (Tuesday, Thursday, Saturday). We chose 6 treatments because in a previous open-label pilot study, which delivered a series of 10 high-dose treatments, 85% of the improvement in depressive severity was observed after the initial 5 treatments [1]. Subjects who were initially randomized to low dose and did not respond after 6 treatments (HDRS-24 improvement < 50% at the Week 2 assessment) had the option to continue with a series of 6 additional open-label treatments using the high-dose intervention.

Two changes were made to the study design part-way through the trial, after the first 7 subjects had been randomized. Under the original design, subjects who did not respond to the first 6 treatments had the option to cross over and receive 6 additional blinded treatments. But it became apparent that any subject who received both types of intervention would be functionally unblinded during the seventh treatment because the recovery experience differed noticeably between low and high dose. Therefore the cross-over component of the design was replaced with an open-label high-dose treatment arm, available to subjects who did not initially respond to the low-dose intervention. This design change did not affect blinding or other trial activities up to and including the primary Week 2 endpoint. The second design change was a broadening of inclusion/exclusion criteria to accelerate enrollment. The age range was expanded from 18-55 to 18-65 years, and the original requirement to document failure of two adequate medication trials was eliminated.

***Participants and baseline assessments***

Treatment-seeking outpatients were recruited from the Treatment Resistant Mood Disorder clinic at the University of Utah, which provides electroconvulsive therapy, transcranial magnetic stimulation therapy, ketamine infusion therapy, and psychopharmacological management. Participants were instructed to continue their current psychiatric medications unchanged for at least two weeks leading up to propofol treatments and throughout the treatment phase. The baseline assessment included a medical history, physical examination, 12-lead electrocardiogram, complete blood count, comprehensive metabolic panel, and pregnancy test (if indicated). Selected modules from the Mini International Neuropsychiatric Interview (M.I.N.I. 7.0.0) were administered by a psychiatrist to confirm the primary diagnosis and characterize psychiatric comorbidities. To evaluate depression severity, the 24-item Hamilton Depression Rating Scale (HDRS-24) was administered using a structured interview [2,3] and the 16-item self-rated Quick Inventory of Depressive Symptomatology (QIDS-SR) was collected [4]. The Montreal Cognitive Assessment (MoCA, version 8.1) was performed to measure baseline neurocognitive function [5]. Subjects completed a series of validated questionnaires to measure depression, anxiety, suicidal ideation, functional impairment, and quality of life: the Patient Health Questionnaire (PHQ-9), the 7-item Generalized Anxiety Disorder scale (GAD-7), the 19-item Beck Scale for Suicide Ideation (BSSI), the 5-item Work and Social Adjustment Scale (WSAS), and the abbreviated World Health Organization Quality of Life scale (WHOQOL-BREF) [6-10].

***Propofol treatments***

Treatments were administered at the Huntsman Mental Health Institute in an interventional suite designed for electroconvulsive therapy. This facility employed standard monitoring as recommended by the American Society of Anesthesiologists: continuous EKG, pulse oximetry, noninvasive blood pressure, temperature, respiratory rate, and capnography. All Covid-19 screening protocols and precautions for aerosol-generating procedures advised by the University of Utah Health System were followed. When subjects arrived in the morning, the study team confirmed fasting for at least 8 hours, measured body weight, reviewed medication changes, and assessed for adverse events. A nurse placed an intravenous (IV) catheter in one upper extremity. A BIS Monitor (BIS VISTA Monitoring System, Aspect Medical Systems) was applied with a 4-electrode sensor (BIS Quatro, Covidien) to measure the left frontal EEG throughout the procedure. The participant was pre-oxygenated, lidocaine was administered IV to prevent pain at the injection site, and ondansetron was given IV to prevent nausea. Other pre-medications were sometimes administered to address side effects (ketorolac, n = 8 subjects; sodium citrate, n = 1; diphenhydramine, n = 1; dexamethasone, n = 1; albuterol, n = 1). An anesthesiologist then administered propofol (2,6-diisopropylphenol; Diprivan injectable emulsion; Fresenius Kabi). A laryngeal mask airway and mechanical ventilation were used for high-dose treatments; endotracheal intubation was an option but proved unnecessary. Airway support and mechanical ventilation were not needed during low-dose treatments. To ameliorate hypotension during high-dose treatments, Trendelenburg positioning was occasionally used, and one participant required pressors (5–10 mg boluses of ephedrine). IV fluids and 100% oxygen were administered throughout low- and high-dose treatments and during recovery. During the recovery phase, additional medication was administered to 4 subjects to address side effects (acetaminophen, ibuprofen, ketorolac, ondansetron, or promethazine). Participants were monitored by a nurse until discharge criteria were met. A responsible adult companion drove the subject home and monitored them for several hours after each treatment.

#### *Outcome assessments*

The primary depression outcome measure was total score on the HDRS-24. The scale was administered by blinded clinical raters (faculty psychiatrists or senior psychiatry residents) who were trained to use the structured interview instrument. Ratings were performed via telephone due to the COVID-19 pandemic. The HDRS-24 was assessed at baseline, Week 1 (after 3 treatments), and Week 2 (after 6 treatments). The pre-specified primary efficacy endpoint was change in HDRS-24 total score from baseline to Week 2. The median time interval from the baseline HDRS-24 to the first treatment was 4 days; the median interval from the sixth treatment to the Week 2 rating was 5 days.

The pre-specified secondary depression outcome was PHQ-9 total score. Other secondary outcomes measured anxiety (GAD-7), suicidal ideation (BSSI), functional impairment (WSAS), quality of life (WHOQOL-BREF), and cognition (MoCA, version 8.2). The PHQ-9, GAD-7, and BSSI were administered at baseline, Week 1, and Week 2; the WSAS, WHOQOL-BREF, and MoCA were administered at baseline and Week 2. MoCA assessments were sometimes not feasible because in-person visits were restricted due to the COVID-19 pandemic.

Assessments during the open-label high-dose treatment phase mirrored those during the randomized phase. The primary and secondary depression endpoints were change from Week 2 to Week 4 in HDRS-24 total score and PHQ-9 total score, respectively. Other secondary outcome measures were the GAD-7, BSSI, WSAS, and WHOQOL-BREF. Before each treatment, a modified Columbia Suicide Severity Rating Scale (C-SSRS) was collected to assess suicidal thoughts over the past 48 hours, and the QIDS-SR was collected (as with ECT patients in our clinic) in order to facilitate subsequent comparison to ECT.

After drug was discontinued, we measured the time to emergence of consciousness. Subjects were considered to meet "eyes-open" criteria when they consistently opened their eyes to verbal command, or when they were able to consistently report their name when asked. Time to re-orientation was measured by asking the date, month, year, day, place, and city repeatedly until all correct answers were obtained (at 3, 5, 10, 15, and 20 minutes after eyes-open). To evaluate the acute subjective effects of propofol with each infusion, the eight PANAS subscales were repeated and the Drug Effects Questionnaire (DEQ-5) was collected post-treatment. During the recovery phase, adverse effects were assessed in five categories (sore throat, nausea, vomiting, discomfort at the IV site, and other side effects).

***Power calculations***

Power simulations during the planning stages were based on the mean HDRS-24 improvement (58%) and variance observed during a pilot study [24]. Those analyses indicated that a sample size of 12 per group would have 87% power to detect a large between-group Hedges effect size of 1.0, corresponding to 58% versus 20% improvement with high- versus low-dose propofol (assuming two-tailed $\alpha = 0.05$). Analyses also showed that a moderate difference (effect size = 0.7; 58% versus 27% improvement) would require 24 subjects per group for 90% power, and a small difference (effect size = 0.4; 58% versus 42% improvement) would require ~100 subjects per group to be adequately powered. Therefore this trial is adequately powered to detect only a large effect size between the low- and high-dose propofol interventions using a conventional two-sided $p < 0.05$ threshold.

***Statistical analyses***

In addition to the primary analysis using a general linear model, pre-planned secondary analyses employed linear mixed models to evaluate whether the rate of change of depression severity (HDRS-24 total score or PHQ-9 total score) differed between low-dose and high-dose treatment groups. The linear mixed model ("lmer" function in "lme4" package, version 1.1-28) was fit using subject intercept as the random effect. The fixed-effect predictors were treatment group, time (baseline, Week 1, Week 2), and group-by-time interaction. Wald $\chi^2$ tests and corresponding p-values were calculated from fitted models ("Anova" function in "car" package, version 3.0-12). A significant group-by-time interaction (two-sided $p < 0.05$) was taken as evidence for a difference between groups in the rate of improvement.

Effect sizes for all outcome measures (standardized mean difference, SMD) and confidence intervals were calculated using the "cohen.d" function with Hedges' correction ("effsize" package, version 0.8.1). A planned analysis of MoCA scores was not performed due to a high proportion of missing data (42%) related to the pandemic.
